# Physical activity and brain health: a systematic review of Mendelian Randomization studies

**DOI:** 10.1101/2025.06.23.25330111

**Authors:** Adrià Túnez, Margot P. van de Weijer, Joseph Firth, Karin J.H. Verweij, Jorien L. Treur

## Abstract

**Objective:** Observational studies consistently link physical activity (PA) to better brain health; psychiatric, neurocognitive, and mental well-being outcomes. While PA intervention studies generally find positive mental health effects, it is unclear whether day-to-day PA, not manipulated through an intervention, is beneficial.

**Design/Data:** Systematic review of Mendelian Randomization (MR) studies using genetic instrumental variables to assess causal effects of PA on brain health. In Embase and medRxiv, 557 articles were identified, of which 35 met inclusion criteria. Study quality was determined based on a MR-specific scoring system.

**Eligibility criteria:** MR studies exploring any PA exposure (self-reported PA, sedentary behaviours, strength-related traits) on mental health, cognition, or cognition-based neurodegenerative outcomes (e.g., Alzheimer’s).

**Results:** Of 35 studies, 43% received a low quality score and 57% a moderate score. There was no consistent evidence for causal effects of day-to-day PA. For sedentary behaviours (e.g., TV-watching) there was consistent evidence for increasing effects on depression. For strength-related traits (e.g., grip-strength) there was consistent evidence for protective effects, particularly on cognitive outcomes. For depression specifically, device-measured PA more often showed protective effects on depression than self-reported PA.

**Conclusion:** MR studies on PA and brain health are generally lacking in quality, due to low sample sizes and/or poorly measured variables. Results are mixed with the most consistent evidence indicating that better physical strength and less sedentary behaviour are beneficial for brain health. Since core MR assumptions are difficult to fulfill for highly complex PA traits, triangulation with other methods and improved phenotyping is needed in future work.

**Summary:** *What is already known?:*

- The prevalence of the most common mental health disorders (i.e., depression, anxiety) has been increasing in the last 20 years in the United States and Europe.
- bservational studies report a consistent association between physical activity and brain health outcomes, but the high risk of confounding precludes a causal interpretation.
- Physical activity intervention studies find positive mental health effects, but do not show whether day-to-day physical activity, not manipulated through an intervention, is beneficial.

*What are the new findings?:*

- verall, the Mendelian randomization literature offers limited support for a broadly protective effect of physical activity.
- Self-reported physical activity showed inconsistent associations with depression, schizophrenia, and bipolar disorder, possibly due to misclassification and confounding.
- behaviours were more consistently associated with negative mental health outcomes, particularly depression.
- Strength-related traits, such as handgrip strength and appendicular lean mass, showed the most consistent and strong, protective effects, especially on cognitive outcomes.
- Randomization relies on core assumptions that are difficult to fulfill for highly complex traits, like physical activity. Triangulation with other methods and improved phenotyping is needed in future work.

## Introduction

In recent years, the term ‘brain health’ was redefined as an umbrella term encompassing all psychiatric, mental well-being and neurocognitive outcomes [1, 2, 3]. Mental health disorders are one component of brain health. They affect almost 30% of individuals across the lifespan [4] and account for 32% of all years lived with disability and 13% of disability-adjusted life years worldwide [5]. The prevalence of the most common mental health disorders (i.e., depression, anxiety) has been increasing in the last 20 years in the United States and Europe [6, 7]. Often, individuals with a mental health disorder also exhibit cognitive dysfunction [8], which can lead to loss of independence, affecting their ability to live a fulfilling life. While treatment and detection are constantly improving, many individuals do not achieve remission with standard treatment [9, 10].

Alongside the development of novel pharmaceutical and therapeutical treatments, the field of psychiatry has placed more focus on daily life activities that influence health, including physical activity (PA) [11, 12]. The World Health Organization (WHO) defines PA as any bodily movement produced by skeletal muscles that requires energy expenditure [13]. According to cross-sectional scientific evidence accrued over the past century, regular PA is positively associated with psychological and physical well-being [14, 15]. Although robust causal evidence is limited, guidelines around PA have already been adopted by several institutions. For instance, both the US Physical Activity Guidelines for Americans [16] and the UK Chief Medical Officers’ Physical Activity Guidelines [17] recommend attaining at least 150 minutes of moderate-to-vigorous PA per week for reducing the risk of depression. Similarly, the WHO currently recommends regular exercise as a means to maintain a healthy cognitive state [18].

The available evidence for causality comes from randomized intervention studies, which show that exercise programs benefit cognitive functioning, depression (severity), sleep quality, and psychiatric symptoms [19, 20, 21]. However, some intervention designs have been identified as low in quality [22], with limitations such as selection bias, attrition bias and inconsistent results in long-term interventions [23]. In addition, it is not certain yet whether daily PA, not manipulated through an intervention, also causally influences mental health or whether people in a healthier mental state feel more inclined or motivated to engage in more exercise.

PA can be measured with a range of measures, which may affect the consistency of results. For instance, self-reported measures can differ greatly from device-based measurements (e.g., reporting exercise times per week vs recorded exercise as obtained from an accelerometer). In a meta-analysis on self-reported PA and mental health, leisure-time and school-based PA were positively associated with better mental health, while work or household-related PA were either negatively associated or showed no association with mental health [24]. In a large-scale cohort study, different patterns of associations between self-reported and device-measured PA and mental health were reported [25]. In summary, many questions remain on the extent to which effects of PA vary across the type and setting [26].

In the past few years, genetically informed research has gained popularity in PA research, providing valuable alternatives to RCTs. A Genome Wide Association Study (GWAS) is used to identify specific genetic variants that are associated with risk for a disease, or any other human trait, with the goal of better understanding its biological underpinnings [27]. Multiple GWASs of PA measures have been published, including device-measured PA, self-reported exercise, handgrip strength, sedentary behaviour, and others. The resulting *summary data* of a GWAS (i.e., the effect estimates summarized across all individuals for all analysed genetic variants) can be used for follow-up analyses, including a causal inference method: Mendelian Randomization (MR). MR uses the random nature of genetic inheritance to study causality in observational epidemiological studies [28]. When certain underlying assumptions hold, confounding and reverse causality are circumvented. MR has been used to assess causal relationships with measures of brain health in multiple studies. However, an overview of the outcomes of such studies, and consistency across different measures of PA, is currently lacking. Therefore, in this study, we review all available MR literature on the effect of PA on different ‘brain health’ outcomes, in order to answer the following questions: (1) Do MR studies find evidence for causal effects of PA on brain health? (2) Are there any differences in results between objectively-measured vs. self-reported PA?

## Methods

### Literature search and review

This study was pre-registered at PROSPERO (CRD42024514302; https://www.crd.york.ac.uk/prospero/display_record.php?RecordID=514302). Available literature was systematically reviewed by searching PubMed and preprint server medRxiv for all MR studies including any PA measure (e.g., exercise, sports, training) as an exposure for causal effects on a range of psychiatric, mental well-being and neurocognitive outcomes (’brain health’). No date restriction was applied. We restricted our search to English-language publications (search strategy provided in Supplementary Material 1). The search was conducted on 24^th^ of June 2024.

Search output was downloaded and uploaded to Rayyan [29], an online tool to organize and simplify literature reviews. Duplicates were identified and excluded. Next, potentially eligible studies were selected to be included based on title and abstract. We followed PRISMA guidelines in extracting and selecting the data. A random subset of 100 studies was selected and screened independently by two reviewers (AT and MW). The selection results were compared and a 97% alignment was achieved (Cohen’s kappa = 0.78). Cases of disagreement were reviewed individually and discussed by the reviewers, and were ascribed to differences in the phenotype inclusion. One of the reviewers was more lenient on the strength-related phenotypes but both reviewers settled for a more strict revision. One researcher (AT) screened the rest of papers.

### Inclusion and exclusion criteria

Any study that included MR analyses exploring the causal effect of any PA exposure(s) on the outcome(s) of interest was included. PA could be measured both through recorded measures (grip strength tests, accelerometer data etc.) and (self-)reported measures (questionnaires, journals etc.). Since the (in)ability to engage in PA is a major determinant of PA-related traits, we additionally included traits like muscle mass or strength, which are often used as proxies for PA among older populations. Measures not exclusively linked to PA, for instance the frailty index (which incorporates several health parameters beyond PA), were excluded.

For the outcomes, all (symptoms of) mental health disorders diagnosed by a professional or measured through self-report on (standardized) questionnaires were included, as well as any cognition-related outcome. Within neurodegenerative outcomes, we differentiated between those primarily associated with cognition (e.g., Alzheimer’s disease (AD), dementia) and those with a predominantly physical component (e.g., ALS, Parkinson’s disease). The former were included whereas the latter were excluded. Papers where PA was only treated as an outcome were excluded. Review papers were excluded.

### Mendelian Randomization

MR is an instrumental variable approach that employs (a set of) genetic variants (most commonly single nucleotide polymorphisms, SNPs) to proxy an exposure and test its effects on an outcome.

Somewhat similar to an RCT, the random segregation of alleles (genes) inherently splits the population into (genetically) ‘exposed’ and ‘controlled’ groups. Because segregation is random, it is unlikely that being a part of one or the other group is influenced by confounding effects that bias conventional epidemiological analyses.

MR relies on 3 core assumptions (Fig. 1): (1) genetic variants used as instrumental variables must be associated with the exposure of interest (relevance); (2) genetic variants used as instrumental variables should be independent from confounding factors (e.g., assortative mating, population structure) also affecting the outcome (independence), and (3) genetic variants do not influence the outcome in any other way than through the exposure (exclusion restriction). Horizontal pleiotropy is when a genetic variant influences the outcome independently of the hypothesised exposure, and it can undermine the assumptions. Various MR sensitivity analyses have been developed to assess potential deviations from these assumptions.

**Figure 1.**
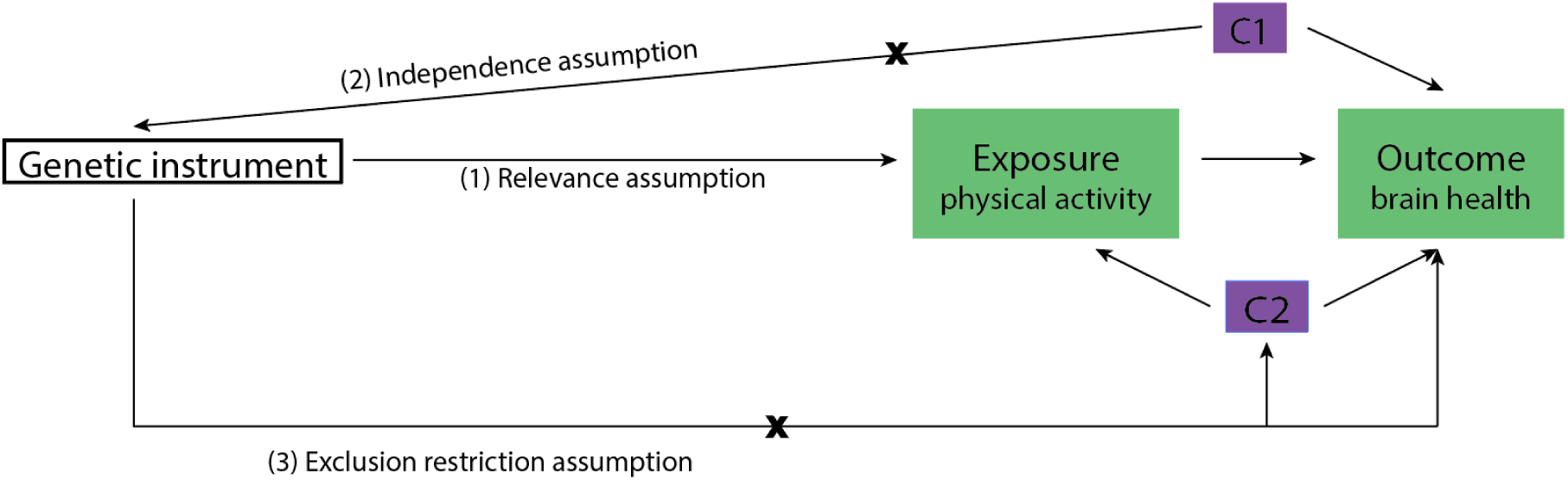
Visual representation of the 3 core assumptions of MR. The genetic instruments used serve as proxies of the exposure -in this example, where physical activity is represented as exposure, the genetic instruments proxy physical activity. C1 refers to confounding mechanisms such as assortative mating and population structure that affect both allele distributions in the population and the outcome and C2 refers to confounders that affect the exposure and the outcome.

‘One-sample MR’ uses individual-level data on the exposure and outcome from the same study sample. ‘Two-sample MR’ derives these associations from two independent GWAS samples instead, allowing large (publicly available) GWAS datasets to be conveniently combined.

### Scoring MR papers

We independently evaluated the quality of the MR papers based on a scoring system developed by Treur et al. 2021 [30] (Supplementary Material 2) which was cross-checked with the most recent MR guidelines [31].

The scoring system is based on several key features: 1) phenotype measurement (including sample size and quality of the exposure and outcome measurements), 2) instrument strength (p-value threshold used to select SNPs, number of SNPs, biological knowledge of genetic variants, F-statistic for instrument strength, percentage of variance that the instrument explains), and 3) sensitivity methods used. Each study was given a final score of ‘–’ (insufficient), ‘– +’ (sufficient) or ‘+’ (good). If both the sample size and the key analytical method requirements were met, the study score was considered sufficient (– + ). When a study had a good sample size and analytical methods and additionally used extensive (sensitivity) methods, a good score (+) was given. We applied a reasonable doubt principle, where we inferred parameters that were not reported but that can be known based on other literature (e.g., if a sample size was not reported but we knew it from other studies or prior research). The selected manuscripts were divided into two sets. Two of three researchers (AT, MW, JT) independently scored each set and later compared the scores. In case of disagreement, all three researchers discussed the differences and reached a final decision.

### Strategy for data synthesis

Our data synthesis strategy was to outline the results, strengths and weaknesses of the included MR studies examining the relation between PA and brain health. We extracted information on the studies’ hypotheses, measure(s) of PA and brain health used, and results of the main MR analysis (one reviewer AT). Additionally, we assessed differences in findings between studies that used objectively measured versus self-reported PA (Supplementary Material 3).

### Equity, diversity, and inclusion statement

The author group is gender balanced and consists of junior, mid-career and senior researchers. However, all members of the author group are from European descent. Our review applied as few exclusions parameters as possible given the scope of the study. Datasets included tend to include mostly European descent individuals given the extended use of UK Biobank in genetics research. No restrictions were applied on age or sex. Possible caveats of the research are disclosed and discussed in the limitations.

## Results

### Searches

A total of 557 articles met our search query, of which 556 were unique (Fig. 2). Of these, 497 were excluded based on title and abstract, mainly due to wrong exposures and outcomes or wrong design (e.g., review or no MR). The remaining 59 articles were retrieved and assessed for eligibility. During screening of the full text, another 24 studies were excluded. Of the final 35 studies included in qualitative synthesis, 13 investigated cognitive outcomes, 20 investigated psychiatric outcomes, and 2 investigated both (see Supplementary Table S1).

**Figure 2.**
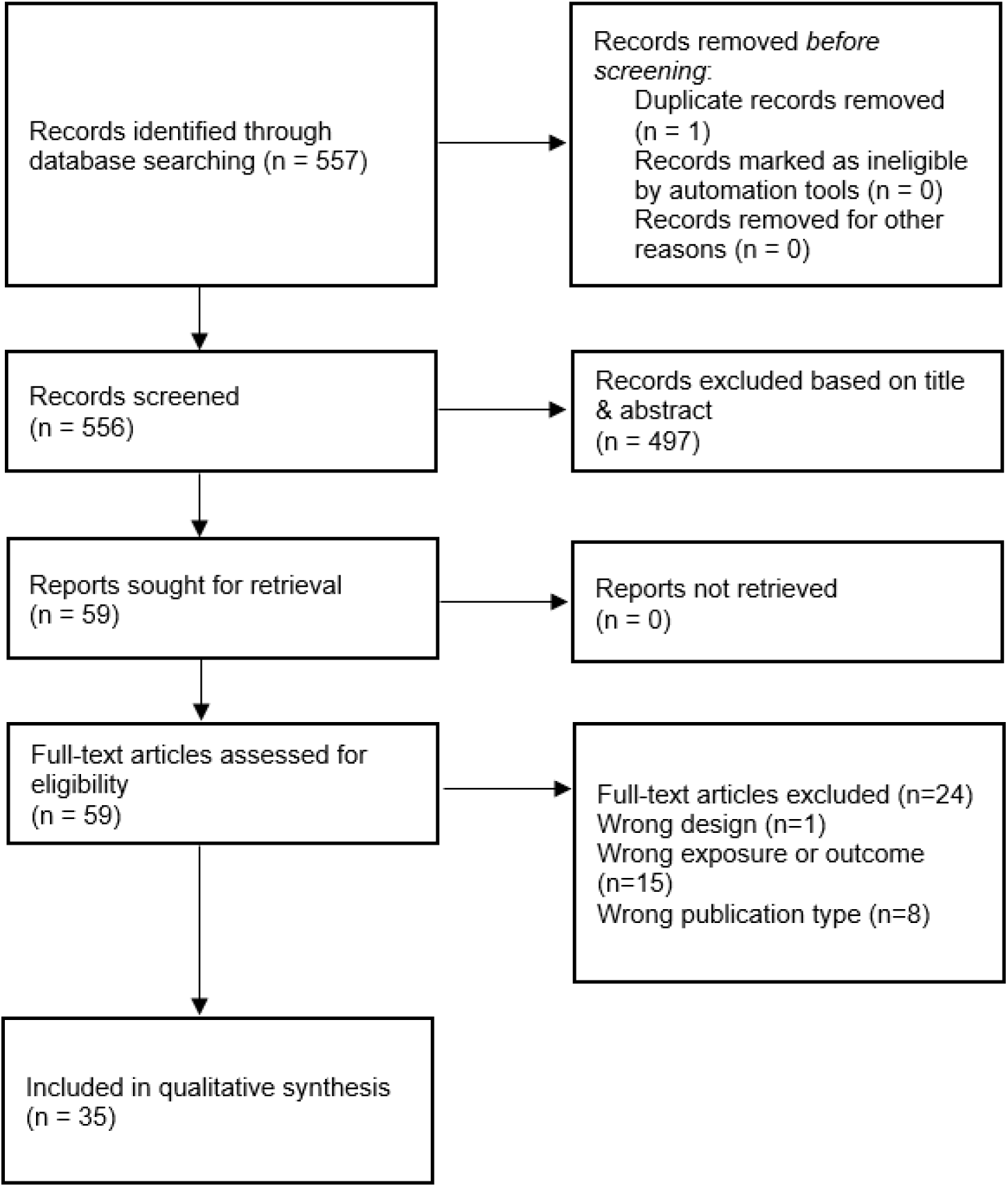
PRISMA flow chart demonstrating the selection of articles included for qualitative synthesis.

### Descriptives

Out of the 35 studies, 15 (43%) received a final score of insufficient (–), 20 a sufficient (-+) score, and none a good (+) score (Supplementary Table S2). While the sample sizes were considered large in 70% of the studies, very few cases (8%) had a sufficiently strong instrument strength (based on a combination of p-value threshold, number of SNPs, biologically informed, F statistics and variance explained reported and sufficient). Mostly due to either relaxed p-value thresholds, small number of SNPs or lack of reported statistics. Only 2 studies presented extensive sensitivity analyses.

Given the limited availability of well-powered PA-related GWASs and brain health measures, 15 studies used the same or partly the same data sets to obtain genetic estimates for both exposures and outcomes. Many of the remaining studies used the same datasets for either the exposure or outcome. In fact, all studies included at least one trait obtained from a GWAS from UK Biobank data, albeit not necessarily from the exact same sample of people. This means that the causal findings presented should not be regarded as (completely) independent (overlap is indicated in Supplementary Table S3).

An overview of the studied exposure – outcome pairs can be seen in the evidence gap map (Fig. 3). Depression, schizophrenia, AD and bipolar disorder were the most studied brain health outcomes, while self-reported PA and accelerometer-measured PA were the most studied exposures. The overall quality of MR studies seems to be higher when self-reported phenotypes are used compared to device-measured phenotypes. This is mostly due to the larger sample size and number of SNPs of self-reported phenotypes.

**Figure 3.**
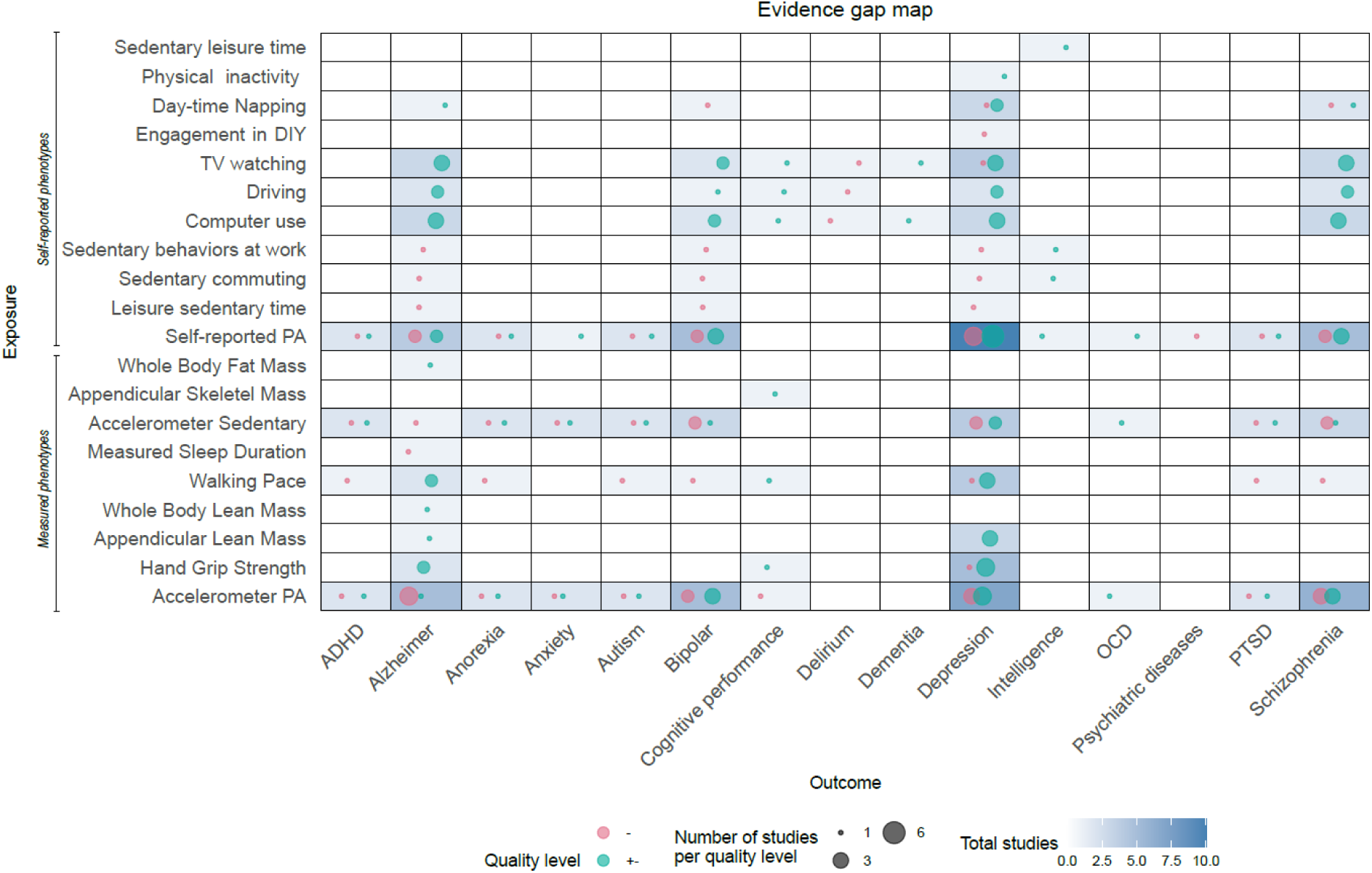
Evidence gap map representing the number of studies (in total and per quality level) for each exposure and outcome of interest. Larger bubbles indicate a larger number of studies, while colour coding reflects study quality (green representing higher-quality and red lower quality studies).

### Effects of device-measured and self-reported PA

#### Mental health traits

There were mixed effects of (accelerometer) measured PA on depression. While most studies [32, 33, 34, 35, 36] reported weak evidence that PA decreases depression risk, some studies reported no effects [37, 38]. The quality of the studies reporting evidence for causality was on average rated lower than those that did not (- vs -+) with results mostly being inconsistent across sensitivity methods. Additionally, Choi et al. (2019) [32] and Iob et al. (2023) [34] evaluated the same exposure/outcome summary statistics. Casanova et al. (2023) [33] and Ba et al. (2024) [35] did too. No evidence for effect of PA on schizophrenia [67, 68, 66, 62] or bipolar disorder was reported [38, 34]. One study reported measured PA decreasing risk of bipolar disorder [40], but its quality was low (-). No effects were found on anxiety or PTSD [38, 34].

There were also mixed effects of self-reported PA on depression risk. While some studies found evidence for protective effects [41, 42, 43], most found no effects [32, 44, 37, 45, 34, 35, 43]. Studies reporting no effect were on average lower in quality than those that did (- vs -+), mostly reflecting lower sample sizes and relaxed p-value thresholds, while the results of those that found effects were consistent across sensitivity methods. No effects were found on schizophrenia [39, 34, 35], bipolar disorder [34, 35], anxiety or PTSD [34].

#### Cognitive traits

Overall, analyses of PA and cognitive traits were more consistent. Several studies reported no effects of (accelerometer) measured PA on AD [46, 47, 48]. Wu et al. (2021) [49] found that PA reduced AD risk in one dataset, but were not able to replicate this results in a validation set. One other study found a protective effect on AD [50], but only when they using a lenient threshold to select instruments. Cheval et al. (2023) [51] reported that measured PA increased cognitive functioning. Liao et al. 2022 [46], Baumeister et al. 2020 [47], Zhang et al. 2022 [48] and Wu et al. 2021 [49] shared exposure/outcome summary statistics.

One study found that self-reported walking reduced the risk of AD [48], while another study found that self-reported moderate PA but not vigorous PA increased the odds of developing AD [52]. No other study found effects of self-reported PA on AD liability [46, 49, 48]. Finally, only one study investigated the effects of self-reported PA on intelligence and found no effects [53]. The quality of all the papers was assessed as low (-). Four studies fully shared exposure/outcome summary statistics [46, 47, 48, 49].

### Effects of device-measured and self-reported sedentary behaviours

#### Mental health traits

There was evidence that sedentary time (accelerometer-measured) increased anxiety and decreased well-being [33] and device-measured sedentary behaviours decreased anorexia and schizophrenia risk [34]. The quality of both papers was assessed as low (-). There was evidence that self-reported TV watching, but not computer use and driving, increased depression risk [54, 35]. The quality of these studies was moderate (-+). Surprisingly, one study found that TV watching protected against bipolar disorder [35]. The quality of this study was moderate (-+).

Negative effects of self-reported sedentary behaviours on depression (sedentary behaviour increased risk of MDD) were found, albeit partially mediated by BMI [41]. After correction for BMI, Wang et al. 2022 [44] found no effect of self-reported leisure sedentary time on depression. Choi et al. 2020 [45] and Chen et al. 2020 [55] found positive causal effects of time spent TV watching and day-time napping on increased depression risk. No effect of self-reported leisure sedentary time was found on MDD, SCZ or BP [37].

#### Cognitive traits

While computer use reduced the risk of AD [54], or had no effect on AD, dementia, or cognitive performance [56, 57], less intensive sedentary activities like TV watching, were reported as detrimental for cognitive performance, dementia, and AD [56, 57]. He et al. 2022 [54] and Yuan et al. 2023 [57] shared exposure/outcomes summary statistics. Zhang et al. 2022 [48] found that neither self-reported nor measured sedentary behaviours were associated with AD. Finally, there was evidence of leisure sedentary time having a negative effect on intelligence while sedentary behaviours at work had positive effects on intelligence. Sedentary commuting had no effect [53].

### Effects of physical strength-related phenotypes

#### Mental health traits

The analyses of strength-related traits on mental health showed more consistent results than the other PA phenotypes. Positive effects of low grip strength on depression were reported by Zhu et al. 2023 [58]. Higher grip strength and walking pace had a protective causal effect on MDD [59, 60]. Zhang et al. 2023 [59] and Wang et al. 2023 [60] shared exposure/outcomes summary statistics. Li et al. 2022 [61] found that left handgrip strength reduced MDD risk while right handgrip strength did not. Lv et al. 2024 [62], however, found that the effect of walking pace and grip strength on depression did not remain significant after correction for BMI. They did find that appendicular lean mass (ALM) remained significantly associated with depression. All papers had a moderate quality rate (-+).

#### Cognitive traits

Walking speed, ASM and handgrip strength were associated with cognitive performance after adjustments [63]. Ye et al. 2023 [64] found that lower ALM and whole body lean mass values increased the risk of AD, whereas Ye et al. 2023 [64] and Jones et al. 2021 [65] found no effect of grip strength on AD. A summarized version of all extracted data can be found in Table 1. Detailed records of all the extracted data can be found in Supplementary Table S4.

**Table 1.**
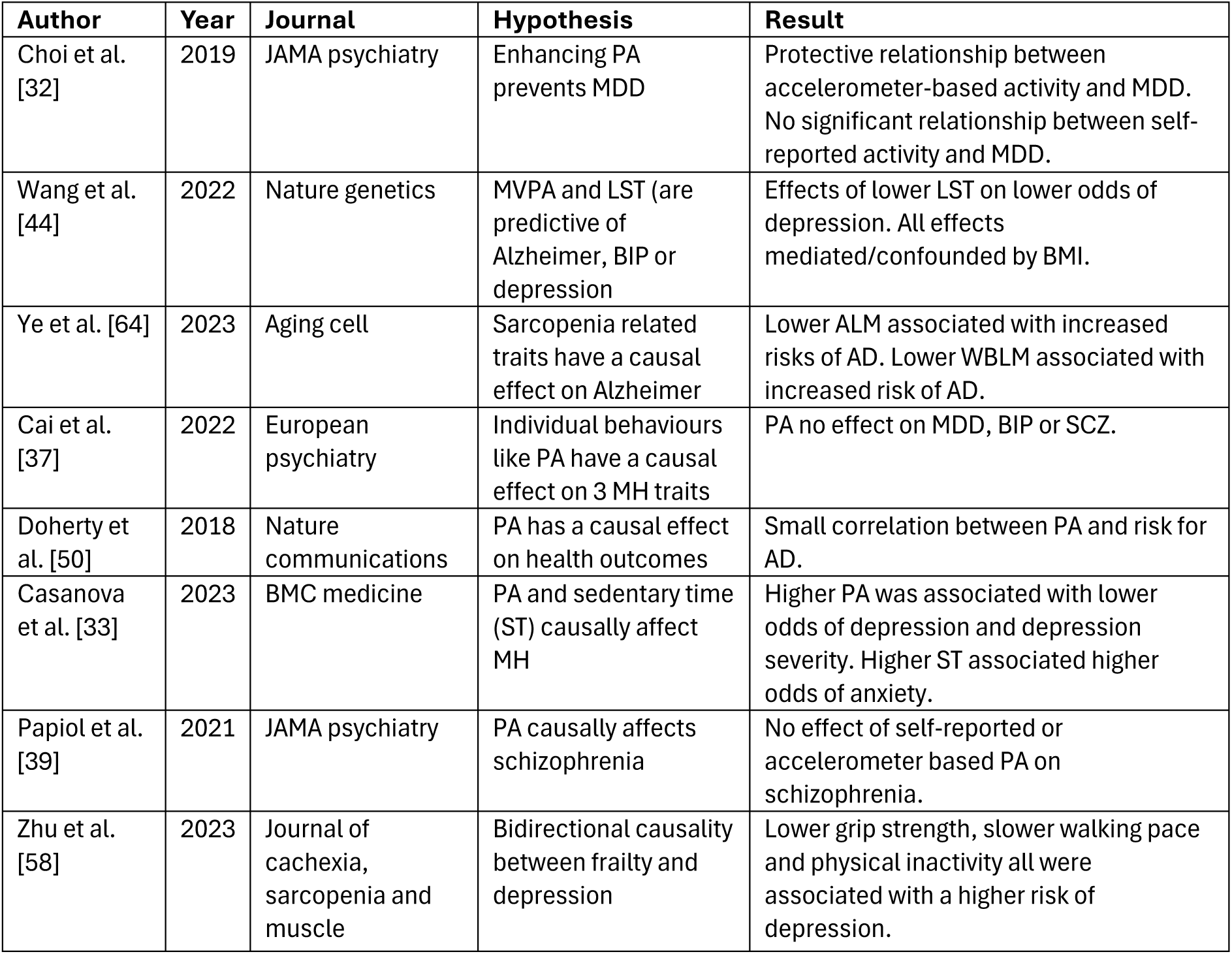
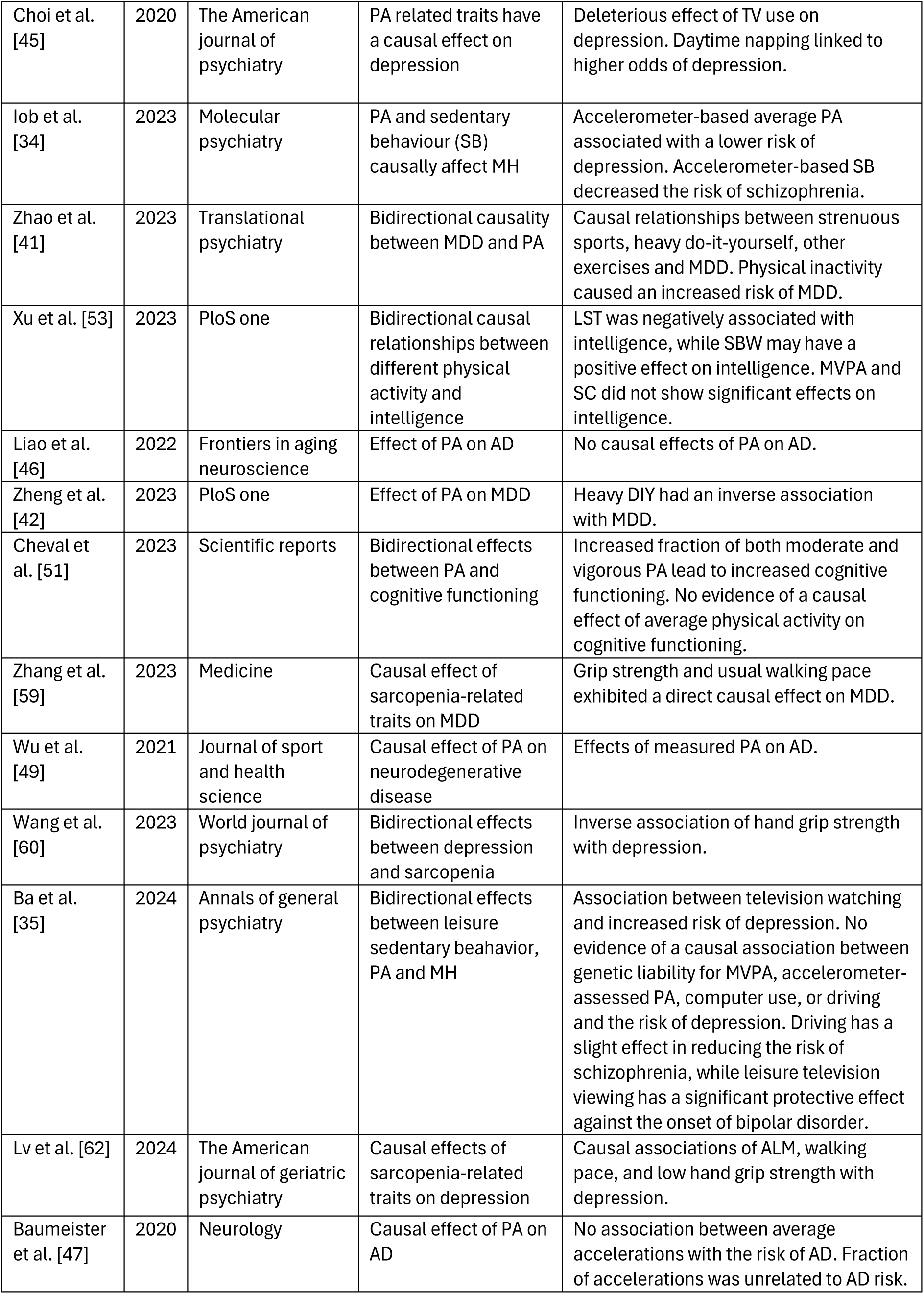
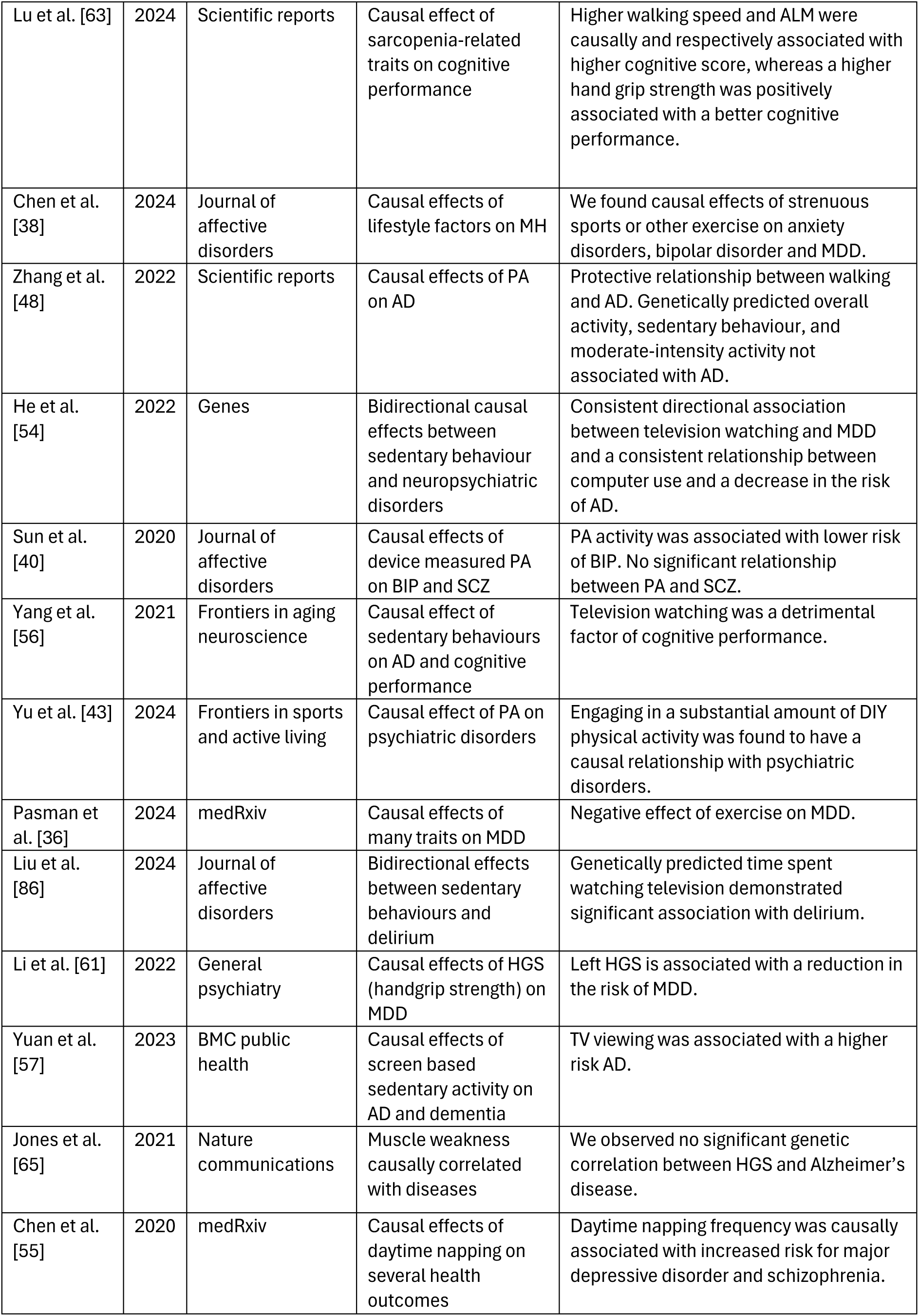
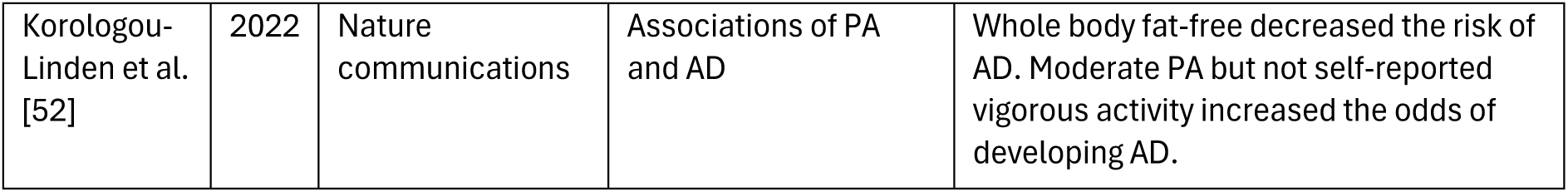
Summary of the studies included and data extracted. Including, from left to right (Author, Year, Journal, Hypothesis, and Result). MDD: Major Depressive Disorder, AD: Alzheimer’s Disease, PA: Physical Activity, MVPA: Moderate to Vigorous Physical Activity, LST: Leisure Sedentary Time, SCZ: Schizophrenia, BIP: Bipolar, ALM: Appendicular Lean Mass, WBLM: whole-body lean mass, SC: Sedentary commuting, SBW: Sedentary behaviour at work.

## Discussion

This review synthesized evidence from 35 Mendelian Randomization (MR) studies examining the potential causal effects of physical activity (PA) and related phenotypes on brain health outcomes. Overall the findings do not offer consistent support for a broadly protective role of PA across outcomes. Rather, the evidence suggests that different types of PA (i.e., being active, sedentary, or strength-related) show distinct patterns of association with brain health.

PA measured as being physically active (e.g., self-reported PA), did not show consistent evidence for protective effects on depression, schizophrenia or bipolar disorder. This contrasts with observational research, which more consistently links PA to reduced risk of these mental health conditions as well as improved cognitive function [21, 66, 67]. This discrepancy may be due to confounding and reverse causation in observational studies, where individuals with better mental health may be more likely to engage in PA. Additionally, self-report PA measures are prone to misclassification and recall bias, potentially diluting true causal effects in MR analyses [68, 69]. These considerations emphasize the importance of using diverse evidence across methods when evaluating the mental health benefits of being physically active.

In contrast, MR studies examining sedentary behaviours such as TV watching or daytime napping found more consistent evidence for negative effects on brain health, particularly increased risk of depression. These findings partially align with observational research showing that prolonged sedentary time is associated with poor mental health and cognitive outcomes [70, 71, 72]. However, caution is still warranted. Sedentary behaviour is closely (genetically) linked to socioeconomic status, physical illness, and other potential confounders that may also influence genetic liability to mental health problems. In MR analyses, this association may further be biased by mechanisms like assortative mating, population stratification, and dynastic effects [73]. Such biases are particularly relevant for lifestyle-related phenotypes like sedentary behaviour, which may be shaped by family environments and social context. To strengthen causal inference in this context, family-based MR designs, which compare siblings or use parental genotypes to control for shared environmental and genetic background, would be especially valuable.

Physical strength traits such as hand-grip strength and appendicular lean mass showed the most consistent evidence for beneficial effects. Although these measures are not strictly PA phenotypes, they can serve as a proxy, are objectively easier to measure than exercise (e.g., amount of muscle mass versus frequency of intense sporting), and are representative of the fitness of individuals that are unable to or do not engage in intense physical activities like running or swimming [74]. Our findings align with recent observational studies [75, 76], though many such studies have focused on older adults [77, 78]. This highlights the need, in future research, to consider age-related comorbidities—such as osteoporosis, arthritis, or cardiovascular disease— which may contribute in both decreasing brain health and physical fitness.

The overall quality of the included MR studies was moderate to low. Of the 35 studies, 20 were rated as “sufficient” and 15 as “low,” with none rated as “good.” These quality assessments highlight the need for cautious interpretation of the findings, especially in light of limitations such as weak instrument strength, potential for horizontal pleiotropy, and inconsistent phenotype definitions across studies.

These caveats may partially explain the discrepancy between intervention studies finding more consistent effects of PA on brain health [79] and MR studies, which seem to be less consistent and generally do not find strong effects. Of special interest are two key differences. First, RCTs are targeted studies that measure the direct, short term impact of an intervention, while MR studies estimate the lifelong influence of genetically predicted exposures. This means that the intervention effects may not be representative of a daily exposure and that the effects might be different due to interventions being short term and MR representing longer term effects. Second, RCTs try to minimize confounding by design while MR relies on strong assumptions about validity of genetic instruments and confounding [28], which may be difficult to meet for complex traits like PA [80]. For example, for traits like alcohol use, there are clear biological mechanisms through which a SNP in an alcohol metabolizing gene affects alcohol use behaviour, but for behaviours like TV watching the mechanism is not clear. A GWAS could capture genetic variants related to traits other than the exposure (i.e., SES), which may affect the target phenotype. This would result in horizontal pleiotropy, violating the exclusion restriction assumption and leading to biased estimates. Such differences highlight the importance of using strict sensitivity methods and integrating evidence across different study designs.

One way to conduct more reliable causal inference is through a process of ‘evidence triangulation’. By combining different methods and data sources, triangulation helps increase the credibility and validity of the results by checking if different approaches lead to the same conclusion [81]. This is important for all MR studies, but especially for those with traits that are highly polygenic and complex (like PA), and where the path between SNP, exposure, and outcome is not completely understood [82]. Supporting the results of this review, a recent triangulation study revealed no effects of self-reported PA on depression/anxiety [83].

Differences between self-reported and device-measured PA likely contribute to the variability in findings that we report. Self-report measures are prone to recall bias and overreporting, though they can capture a broader range of behaviours over longer periods. Device-based measures are more precise but focus primarily on locomotor activity, potentially missing non-aerobic or resistance-based exercise [68, 69]. A genetic correlation of 0.49 has been reported between self-reported and device-measured PA GWASs [25], indicating partial but incomplete overlap. In our review, protective effects on depression were more often found in MR studies using device-measured PA. However, these studies had smaller sample sizes, limiting statistical power. It has also been suggested that current mood and cognitive biases may influence self-reported activity more than objective measures [84]. In future research, studies should use both types of measures and apply stringent sensitivity analyses to improve the reliability of genetic instruments.

This study is not exempt of limitations. First, although we focused on PA as an exposure, many related (accelerometer PA, physical inactivity etc.) and proxy-phenotypes (handgrip strength, lean muscle mass etc.) are included, resulting in a large amount of exposure phenotypes in a limited number of studies. While this reflects the complex nature of PA, finding systematic ways of grouping and assessing phenotypes would be desirable in future research, ensuring stronger, more reliable genetic instruments. Second, most studies relied on GWASs from similar samples, particularly the UK Biobank, which limits generalizability. UK Biobank participants are predominantly of European ancestry, older, wealthier, healthier, and more educated than the general population, with lower rates of smoking, obesity, and chronic disease [85].

In conclusion, this review of MR studies supports a protective role of strength-related traits on cognitive outcomes and negative effects of sedentary behaviours on depression. However, no clear support was found for causal effects of general physical activity on mental health outcomes, regardless of whether activity was self-reported or device-measured. We conclude that using MR as a standalone tool for causal inference is far from optimal for complex traits where there is no clear biological pathway between the included genetic variants and exposure of interest. Despite the numerous advantages of MR, the difficulty to obtain reliable biologically-informed instruments is one of the current challenges in the field. We therefore urge researchers to combine MR with observational, interventional, and family-based designs, while prioritizing consistent phenotype definitions and robust sensitivity analyses to strengthen causal inference.

## Data Availability

All data produced in the present work are contained in the manuscript

## Data availability

Tables summarizing the data extracted are available in the Supplementary tables. GWAS summary data is available in the original study papers.

