## Supplementary Materials for "Physical activity and brain health: a systematic review of Mendelian Randomization studies"

**Content**

1.Search query	2

2.Mendelian Randomization scoring system	3-5

3.Data extraction parameters	6

**1. Search query**

Physical activity:

((physical activity[Title/Abstract]) OR (physical activity[MeSH Terms]) OR (exercis*[Title/Abstract]) OR (exercis*[MeSH Terms]) OR (sport*[Title/Abstract]) OR (sport*[MeSH Terms]) OR (walking[Title/Abstract]) OR (walking[MeSH Terms]) OR (intensity activity[Title/Abstract]) OR (intensity activity[MeSH Terms]) OR (resistance training[Title/Abstract]) OR (resistance training[MeSH Terms]) OR (muscle[Title/Abstract]) OR (muscle[MeSH Terms]) OR (sedentary[Title/Abstract]) OR (sedentary[MeSH Terms]) OR (screen time[Title/Abstract]) OR (screen time[MeSH Terms]) OR (screentime[Title/Abstract]) OR (screentime[MeSH Terms]) OR (aerobic[Title/Abstract]) OR (aerobic[MeSH Terms]) OR (fitness[Title/Abstract]) OR (fitness[MeSH Terms]) OR (chronic exercise[Title/Abstract]) OR (chronic exercise[MeSH Terms]) OR (regular exercise[Title/Abstract]) OR (regular exercise[MeSH Terms]))

Mendelian Randomization

((Mendel*[Title/Abstract]) OR (Mendel*[MeSH Terms]) OR (MR[Title/Abstract]) OR (MR[MeSH Terms])) AND (Mendel* random*)

**2. MR Scoring system from Treur et al. 2021**

**Supplementary Material 2.** Scoring system to appraise the quality of the included Mendelian randomization (MR) studies

| **Type of method** |  | **-** | **- +** | **+** | **Notes** |
| --- | --- | --- | --- | --- | --- |
| **all** | Phenotype measurement* |  |  |  |  |
|  | **a**. Sample size exposure | <50,000 | 50,000 – 100,000 | >100,000 | *Effective* sample size for case-control studies |
|  | **b**. Sample size outcome | <50,000 | 50,000 – 100,000 | >100,000 | *Effective* sample size for case-control studies |
|  | **c**. Type of measure for exposure | Single or a few (survey) question(s) | Validated, multi-item survey questionnaire | Extensive (cognitive) measure in clinic / clinical diagnosis |  |
|  | **d**. Type of measure for outcome | Single or a few (survey) question(s) | Validated, multi-item survey questionnaire | Extensive (cognitive) measure in clinic / clinical diagnosis |  |
|  | Instrument strength** |  |  |  |  |
|  | **a**. *p*-value threshold | ≥5E-08 | <5E-08 | - |  |
|  | **b**. # SNPs included | <3 | 3-10 | >10 | Note that **b** concerns the genetic instruments that include SNPs below the *p-*­value threshold <5E-08 (and *not* additional instruments that include less-significant SNPs ≥5E-08) |
|  | **c**. Biological knowledge of genetic variants | Limited | Reasonable | Good |  |
|  | **d**. F statistic reported | No | Yes | - |  |
|  | **e**. F statistic sufficient | <10 | ≥10 | - |  |
|  | **f**. % variance explained reported | No | Yes | - |  |
|  | **g**. % variance explained sufficient | Low (<1%) | Moderate (1-3%) | High (≥3%) |  |
|  | Bidirectional effects tested | No | Yes | - | n.a. when bidirectional causality is not plausible |
| **One-sample MR (individual level data)** | Main analysis | Regression analysis (SNP-outcome association) | 2-stage least-squares regression (2SLS) | - |  |
|  | Type of genetic instrument | Unweighted allelic score | Weighted allelic score | - |  |
|  | Sensitivity analyses |  |  |  |  |
|  | **a**. Method addressing horizontal pleiotropy | No | Yes | - | E.g. testing associations between instrument & potential confounders |
|  | **b**. Leave-one-out analysis or forest plot of  individual SNP-effects | No | Yes | - |  |
|  | **c**. Considerable extra effort with additional  sensitivity method(s) | No | - | Yes | E.g. using negative / positive controls or directly comparing findings to another, *non*-MR research method (triangulation) |
| **Two-sample MR (summary level data)** | Appropriate temporality | No | Yes | - | If trait 1 is measured in childhood GWAS and trait 2 in adulthood GWAS, then causality from trait 2 🡪 trait 1 can’t logically be tested |
|  | Mention of harmonization of genetic variants across the datasets | No | Yes |  | Should be scored as ‘-+’ if harmonization is explicitly mentioned by the authors or when MR Base was used |
|  | Estimates SNP-exposure and SNP-outcome are from the same ethnic group | No | Yes | - |  |
|  | Sample overlap |  |  |  |  |
|  | **a**. Is % overlap reported | No | Yes | - |  |
|  | **b**. Is there sample overlap | Yes | No | - | Should be considered especially problematic if sample overlap is high |
|  | Sensitivity analyses |  |  |  |  |
|  | **a**. Addressing horizontal pleiotropy | No | Yes | - | E.g. MR-Egger which explicitly estimates horizontal pleiotropy or an approach that excludes or down-weights outlier SNPs |
|  | **b**. Leave-one-out analysis or forest plot of  individual SNP-effects | No | Yes | - |  |
|  | **c**. Considerable extra effort with additional  sensitivity method(s) | No | - | Yes | E.g. using negative / positive controls, directly comparing findings to another, non-MR research method (triangulation), or using many different MR sensitivity methods with contrasting underlying assumptions |

SNP = Single Nucleotide Polymorphism, GWAS = Genome-Wide Association Study. Note: where absolute thresholds are used to judge the quality of a particular aspect of the study (e.g. sample size), it should be noted that these are somewhat arbitrary and were merely used to provide an indication of quality. The formula used to calculate the effective sample size for case-control studies = *4 / (1 / ncases + 1 / ncontrols).* Regarding sample overlap, note that we only indicate whether or not there is (some) overlap between samples, and not the amount, given that there is no reliable threshold from which to infer the potential effects of overlap on causal estimates. In addition, the percentage of overlap should be taken with respect to the larger dataset (e.g., if the smaller dataset has 1,000 individuals and all of these individuals are also in the larger dataset of 10,000 individuals, sample overlap is 10%) as indicated by Burgess et al., 2016, *Genetic Epidemiology*.*With regards to ‘Phenotype measurement’, a very well measured phenotype in a moderate sample size may be just as powerful as a more superficially measured phenotype in a very large sample. However, in case of very small sample sizes (e.g. n = 180 such as in the study by Irons et al., 2007) even an extremely thoroughly measured phenotype will not lead to a high total score. **With regards to ‘Instrument strength’, when a study uses a single genetic variant that explains a relatively large amount of the variance and for which there is good biological knowledge, the fact that only 1 SNP was used is not necessarily problematic. For example, this is the case for SNP rs1051730 in the nicotinic acetylcholine receptor *CHRNA5/A3/B4* gene cluster – each additional risk allele increases smoking heaviness with 1 additional cigarette smoked per day (Katikireddi SV et al., 2018).

**3. Data extraction parameters**

Below we listed all data that was extracted from the full text of the final selection of to be included studies.

- Title, author(s), publication year, journal name.

- Hypothesis

- Study methods details:

o Participants characteristics (age, sex, inclusion criteria, ethnicity) (sometimes not reported in MR studies but available in the original GWAS papers)

o Type of Mendelian randomization design (one-sample or two-sample)

o In case of two-sample MR; whether or not there was sample overlap

o Measure(s) of physical activity and neurocognitive functioning/psychiatric disorder used

o Genetic variant selection (including a description of the GWAS sample that was used to identify the genetic instrument and effect estimates, covariates included, p-value threshold used for genetic instrument selection, whether or not proxy SNPs were used and under which LD threshold, if available a list of the single nucleotide polymorphisms included in the genetic instrument/proxy).

o Any information on statistical power (including % of variance that the genetic instruments explained)

o Any information on multiple testing and correction(s) for it

o Results of the main Mendelian randomization analysis; measure of effect size and precision (e.g., 95% confidence interval).

o Sensitivity analyses to determine robustness of the main finding

o Statistical program used for analysis

o Discussion points/limitations regarding the results
